# Improving Predictive Models With Causal Methods – Study Protocol

**DOI:** 10.1101/2025.03.31.25325004

**Authors:** Yoel Mittelberg, David Rawlinson, Daniel Stiglitz, Gideon Kowadlo

## Abstract

**Background:** In a previous study, Kowadlo et al. [1] developed algorithms (POP – Patient OPtimizer) to predict outcomes for surgical patients at Austin Health. The POP algorithms predict postoperative complications, kidney failure, and hospital length-of-stay. The findings highlight the potential of risk prediction to improve health outcomes and justify further work to improve performance and generalisability.

**Objectives:** The objectives are to:

- Establish and validate a causal graph of elective surgery in a hospital setting
- Test whether causal inference can be used in algorithm development to improve generalisation of predictive models to different patient cohorts
- Implement and test the concept of ‘preventable risk’, risk stratification that combines risk prediction with causal effect; to assist in decision making

**Method:** In order to achieve the objectives, we will:

1. Apply causal discovery methods to the INSPIRE dataset (Lim et al. [2]), to create a causal graph
2. Validate the graph by combining clinical input with data analysis, to identify relevant confounding and collider variables
3. Methodically control for confounders and colliders, while training and evaluating predictive models for length-of-stay, mortality, readmission or complications
4. Measure the generalisation of predictive models across patient populations, when controlling for identified confounders and colliders
5. Implement Conditional Average Treatment Estimate (CATE) and combine it with risk prediction to calculate preventable risk

## 1 Introduction

In a previous study, Kowadlo et al. [1] developed novel perioperative optimiser (POP) algorithms to predict outcomes for surgical patients at Austin Health with a dataset of 11,475 adult admissions. The POP algorithms successfully predicted postoperative complications, kidney failure, and hospital length-of-stay (LOS) with high accuracy, but had lower accuracy for the rare events of readmission and mortality. The findings highlight the potential of risk prediction to improve health outcomes.

In 2024, a public dataset called INSPIRE Lim et al. [2] was released. It contains approximately 100,000 patients and over 126,000 surgical admissions. It provides an opportunity not only to validate the algorithms developed in POP but also to expand the methodology to improve generalisation.

### 1.1 Causal analysis to improve generalisation

Risk prediction algorithms can improve patient care and operational efficiency, but their performance depends on the data used for training. Hospitals serve different populations and vary in geography, demographics, socioeconomics, and clinical characteristics; models developed at one hospital may not generalise to another– even if their clinical processes are similar. Even within one hospital, patient demographics, resources and hospital policies may change over time. This is known as ‘dataset shift’ [3]. A model that was trained at a particular point in time may rely on patterns in the data that no longer exist when it is used to predict risk.

Causal relationships describe the processes that generated a given dataset, which in health typically reflect biological or medical processes related to health conditions and their treatments. In addition to describing these relationships, hospital data may include variables that are coincidentally correlated with the outcome. For example, a hospital may assign patients to a specific ward when the likelihood of surgical success is high. A purely predictive model may learn this association and predict surgical risk based on the ward assignment, rather than the underlying conditions that led to the assignment. If the model is then used in a different hospital, which does not assign wards in the same way, it will fail. By understanding the causal relationships, variables that introduce bias can be excluded or controlled so that the model relies on more general features and suffers from less bias in different environments.

Previous research suggested that causal relationships between variables could induce bias in trained models. Hernan and Robins [4] discuss the impact of confounders and colliders on causal inference and how improper variable selection or conditioning can lead to bias. Arjovsky et al. [5] describe machine learning challenges that are caused by selection bias and confounding factors. They differentiate between spurious correlations and those that are invariant across ‘environments’, which they describe as circumstances such as locations, times and experimental conditions. Schölkopf et al. [6] focus on environments that result in distribution shifts and make the argument that identifying the data’s underlying causal model should improve generalisation ‘out-of-distribution’.

These insights are applicable to most ML models, including our model developed in the previous POP study, which use available features without taking into account causal relationships underlying the data. While the resulting model performed well on the given dataset, it may have included bias that would adversely affect performance on out-of-distribution data, such as future samples (due to dataset shift) or specific sub-populations.

In the context of decision support, any clinical choice that can be made when preparing for, during, or after surgery, can be considered a treatment that may impact the outcome. Even in the context of general risk prediction, i.e. without a prescribed treatment choice, the causal relationships between covariates and the outcome are relevant.

As part of causal discovery, this study will create a causal graph to describe and validate causal assumptions with clinical experts, distinguishing true relationships from spurious ones while validating that clinical expectations are reflected in the data. By guiding which variables to control, this approach should reduce collider and confounder bias, and result in a more generalised predictive model. The model may perform slightly worse on the population that was used to train it, if variables that include some predictive signal are excluded, but perform better on other (out-of-distribution) populations.

### 1.2 Preventable risk

Once a causal model is developed for a predictive scenario, it could be used to provide clinicians with additional insight. For example, to identify patients for whom interventions are most relevant and meaningful [7, 8].

In the context of hospital surgery, causal analysis could identify patients whose risk of complications or prolonged LOS could be mitigated by taking particular steps, such as prehabilitation to reduce weight/body-mass-index (BMI), using a particular type of anaesthetic, or following a specific protocol. This would not only improve patient outcomes, but could eliminate the use of unnecessary resources.

We propose combining a predictive model, where causal bias is controlled, with conditional treatment effect estimates, to identify ‘preventable risk’, i.e. risk that is high and can be addressed clinically. To simulate the clinical setting, choice of ‘treatment’ will be limited to variables that clinicians can modify, such as anaesthetic type, surgical procedure or medication.

Preventable risk will be calculated as a product of risk prediction and conditional treatment effect estimate. Patients who receive high preventable risk estimates would be those who have both a high risk and modifiability of risk factors, allowing clinicians to prioritise patients for treatment based on potential impact.

### 1.3 Impact of study

The study could impact several areas of clinical practice and research:

1. Making predictive models more generalisable would increase the applicability of developed models across hospitals and over time within a hospital, and reduce the complexity of implementing machine learning by reducing the need to train local models.
2. The paper will illustrate the benefit of creating an explicit causal graph, a process that researchers and clinicians could use to reflect on their assumptions about data generating processes in the hospital setting. It could identify areas in which the data does not align with clinician expectations, which may indicate issues in data collection, analysis, or understanding of the underlying causal processes. Insights could assist the improvement of clinical practice, as well as advance research into causal discovery and inference methods.
3. The confirmation that causal methods can improve generalisability of predictive models could stimulate their adoption as part of machine learning methodology.

## 2 Method

We will evaluate the method with multiple model types, such as logistic regression (LR) (using scikit-learn) and extreme gradient-boosted decision trees (XGBoost, using the XGBoost package [9]). Models will be developed and cross-evaluated on two populations (i.e. developed on one population and evaluated on the other, and vice versa) to evaluate generalisation. The method will consist of six main stages, as illustrated in Figure 1:

**Figure 1:**
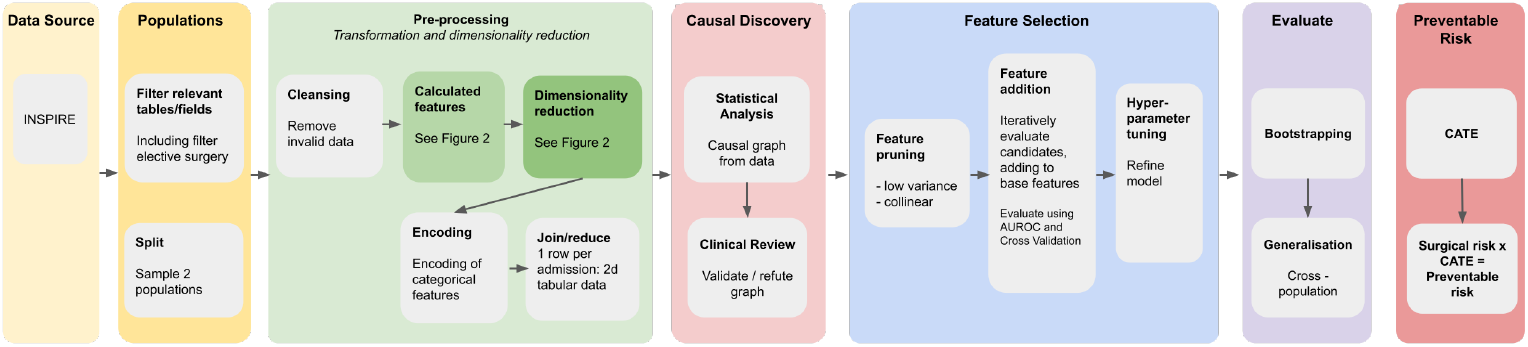
Data processing pipeline

1. The data will be split into two sub-populations
2. Each sub-population will be pre-processed to reduce dimensionality and transform relational data into a tabular format.
3. Causal analysis will map the causal graph in one population and control for bias.
4. Features will be selected to optimise prediction scores on one population.
5. Model performance and generalisation will be evaluated on both populations
6. Preventable risk will be calculated as a product of surgical risk and CATE.

### 2.1 Cohort selection

Inclusion criteria:

- Age: 18 years and older
- Patients undergoing elective surgery

### 2.2 Data source

We will use INSPIRE [2], a dataset published in 2024. It contains more than 126,000 surgical admissions from approximately 100,000 surgical patients in Seoul National University Hospital, South Korea. The data are de-identified and contain information about procedures, lab tests, medications, and diagnosis. It also includes vital signs, which will not be used, but would likely provide additional useful information for predictive modelling.

### 2.3 Creating sub-populations

Given that the study uses one dataset and the objective is to test generalisation across different populations, the dataset will be split into two populations (subsets) in a way that aims to:

1. Maintain the same causal relationships within both populations.
2. Introduce differences between populations which would bias a predictive model trained on one population as compared to the other.

Although BMI is expected to have clinical significance, it can be used to split the population in a way that resembles real-world examples of hospital populations documented in the research literature:

1. Knoedler et al. [10] describe drift in BMI over time, based on the ACS-NSQIP database (2008-2020). During the review period, median age increased by 3 years and median BMI by 0.9 kg/m^2^.
2. Eum et al. [11] compared BMI distribution between patients in the USA (multiple ethnic groups) and Korea. They found different distributions between the countries as well as some differences between ethnic groups in the USA. Example populations are listed in Table 1

**Table 1:** Mean BMI in different populations.

|  | USA |  |  | Korea |
| --- | --- | --- | --- | --- |
|  | Mexican<br>American | Non-Hispanic<br>White | Non-Hispanic<br>Asian | Korean |
| BMI (kg/m<sup>2</sup>, mean ± SD) | 30.89 ± 0.21 | 29.75 ± 0.17 | 25.56 ± 0.14 | 24.04 ± 0.03 |

Based on the studies cited above, the simulated populations will be generated to target a difference of 5 kg/m^2^ between their means (similar to the difference between Korean and non-Hispanic Asian populations, approximately 24-25 and White or Mexican Americas, approximately 29-30).

### 2.4 Pre-processing

Pre-processing will consist of several steps.

#### 2.4.1 Admissions

1. Weight and height will be validated against acceptable ranges and admissions with invalid values will be removed.
2. Length-of-stay (LOS) will be calculated as the time between admission and discharge, in hours.
3. Categorical variables (sex, race, ICD-10 procedure code, emergency/elective, department, anaesthetic type) will be one-hot encoded. Patient variables will be aggregated as the first value in each admission, and procedure variables will be quantified by counting the occurrences of each value per admission.
4. Combining admission procedures: each admission may involve multiple procedures, identified by ICD-10-PCD codes, without explicit indication of the primary surgery, which is the reason for admission. To address this, all procedures will be grouped using the admission ID. The primary operation will be identified as the procedure with the longest operating room duration. Other procedures performed during the same admission will be quantified by counting the occurrences of each procedure type.
5. To make the variables human-readable and interpretable, the name of each admission’s primary procedure will be determined using a spreadsheet from the Healthcare Cost and Utilization Project (HCUP) [12].
6. The outcome of 30-day readmission will be calculated using admission and discharge times

#### 2.4.2 Clinical indicators

Lab tests, diagnosis, and medications will be processed in a similar way:

1. Entries will be matched to a hospital admission if the entry timestamp falls between the admission start and discharge dates.
2. Categorical variables in the entries will be one-hot encoded and split into two groups–those done before the main procedure and those done after.

For lab tests, additional features will be created for the latest result of each test type during the admission and within 6 months prior to surgery. For medications, features will be created for the medication’s therapeutic class, which will be identified by using the first 3 characters of its ATC code. For diagnosis, codes will be grouped according to the ICD-10 ranges, as listed by the CDC table index 2025 [13].

#### 2.4.3 Merge data by admission

Pre-processed data of admissions, lab tests, medications and diagnoses will be joined using the admission ID. The result will be a dataset with one row per admission, on which predictive models can be trained. The process is illustrated in Figure 2.

**Figure 2:**
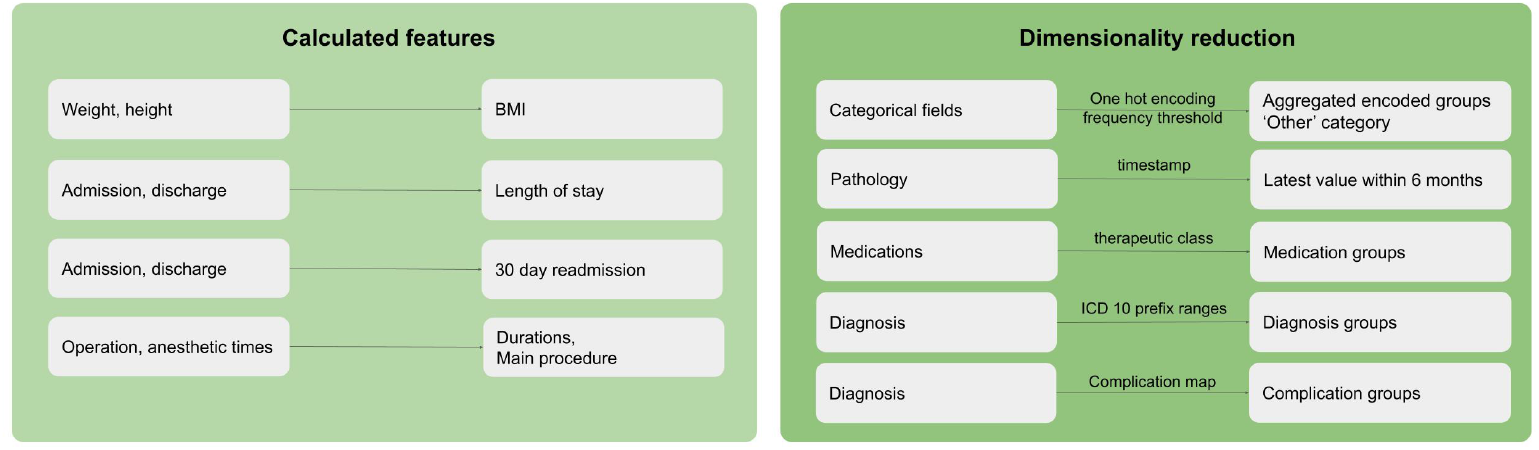
Dimensionality reduction and calculated features

#### 2.4.4 Missing data and class balancing

Missing categorical data will be treated as legitimate input by creating a ‘missing’ category. During model training, missing numerical data will be imputed with XGBoost’s in-built mechanism and by using the median for LR. For LR, numerical data will be standardised (using scikit-learn’s StandardScaler). Class balancing will be achieved by applying a higher weight to under-represented classes.

### 2.5 Predicted outcome

Prolonged LOS will be calculated on a procedure-type basis, by comparing each admission’s LOS with the 75th percentile LOS of the relevant procedure. It is a binary outcome – ‘prolonged’ or ‘not prolonged’.

### 2.6 Causal analysis

Typically, casual analysis seeks to identify causal relationships in datasets and estimate the causal effects of specific treatments. This can be used to define treatment policies to maximise desired outcomes. Dataset variables are typically divided into 3 categories:

1. Outcome: the measure that is being predicted or explained. For the purpose of this study, the outcome of the causal inference will be the same as the outcome of the predictive model.
2. Treatment: a variable representing an intervention or exposure, the impact of which on the outcome is the focus of investigation.
3. Covariates: other variables in the dataset that may have some influence on the outcome and/or the treatment.

#### 2.6.1 Causal discovery

Causal discovery will be used to construct a causal graph from the pre-processed variables. The Peter-Clark (PC) algorithm [14], implemented in the causal-learn Python library [15], will be used for causal discovery to construct a causal graph. The graph will then be validated and refuted using the DoWhy package [16] to ensure that the assumptions align with the data. Additionally, the Fast Causal Inference (FCI) algorithm [17] will be employed to gain insights into hidden confounders that may not be captured by the PC algorithm. This combined approach will leverage the strengths of both methods to better understand causal relationships and address potential biases in the data.

The result of this process will be a visualisation of causal relationships between variables, reflecting statistical relationships in the data as well as expert knowledge.

#### 2.6.2 Mitigation of bias from colliders and confounders

The causal graph will be used to identify colliders and confounders, with BMI as the treatment and prolonged LOS as the outcome. Multiple controls [4] will be tested, including:

1. Inverse probability weighting
2. Standardisation
3. Stratification
4. Matching

The impact of these controls will be evaluated in several ways using DoWhy [16]:

1. Comparison of treatment estimates from different adjustment methods–consistent results would indicate more reliable adjustment.
2. Sensitivity analysis to assess impact of unmeasured confounders, by introducing a synthetic confounder and measuring its effect. If the effect is small, it indicates that the model is robust to unobserved confounders.
3. Negative control analysis to detect residual bias, for example, changing the data in a way that should not change the estimate (such as subsample, or random common cause), or changing the data in a way that should result in a causal estimate of 0, such as placebo treatment or dummy outcome.

The resulting variables from this phase will be used as available features in feature selection.

### 2.7 Feature selection

Variables with very low variance, defined as sufficiently high ratio between the highest occurring value and the second highest, will be removed.

Feature selection will start with a base model that includes basic patient variables. The algorithm will then incrementally add features. In each iteration, every available feature will be tested by adding it to the model and evaluating its performance using the area under the receiver operating characteristic curve (AUROC) with 10-fold cross-validation. This process will continue until all features are tested. Each combination is a ‘feature set’, and the feature set that resulted in the highest score will be selected as the chosen set of features.

Hyperparameter tuning will then be used to optimise results.

### 2.8 Evaluation

Performance will be evaluated from several perspectives:

#### 2.8.1 In-population model evaluation

Within each population, a predictive model will be trained on a subset of the data using logistic regression and XGBoost. Performance of the model will be scored using AUROC and confidence intervals will be calculated with non-parametric bootstrapping using 1,000 iterations. AUROC is a standard metric and is most common in the related literature.

#### 2.8.2 Generalisation

In addition to scoring the models on the population that they were trained on, each model will be scored on the other population to test its ability to generalise.

‘Generalisation’ will be measured by comparing the mean score of both models, before and after applying causal analysis, on the respective populations on-which they were *not* trained. An improvement in mean score would indicate improved generalisation.

### 2.9 Feature importance

Feature importance of the models will be inspected to gain insight into the effectiveness of causal analysis. After applying causal analysis, we expect feature importance to align with causal features rather than purely predictive ones. This alignment should improve the model’s ability to generalise across both populations, as causal features are less likely to be dependent on correlations that exist in only one population.

### 2.10 Preventable risk

Preventable Risk will be calculated as the product of surgical risk and the Conditional Average Treatment Effect (CATE) of modifiable features, expressed as: Preventable Risk = Surgical Risk × CATE.

For example, if two patients have the same surgical risk, but one has a high CATE of modifiable features while the other has a low CATE, the patient with the higher CATE will have a higher preventable risk score, indicating greater potential for risk reduction through clinical intervention.

To calculate preventable risk, the following inputs will be used:

- Surgical risk: determined by the predictive model developed using causal analysis in step 2.7.
- CATE: calculated for multivariate treatments identified by subject matter experts as modifiable by clinicians.

## 3 Related work

Causal Inference has begun to gain attention in healthcare [18, 19, 20, 21] and Causal Machine Learning has been applied in perioperative medicine to enhance risk prediction and optimise treatment decisions. We surveyed the surgical casual ML literature and summarised the studies below.

Lee et al. 2022 [22] estimated causal effects and compared to associative factors (linear regression) for LOS. The patient cohort consisted of cardiac surgical patients undergoing isolated coronary artery bypass grafting or aortic valve replacement surgery. The researchers learnt the causal graph using fast causal inference (FCI), then estimated causal effects using the LV-IDA algorithm [23]. They compared that model with linear regression for LOS to contrast statistical and causal associations. They found that clinicians’ beliefs about causes were mainly correct (some direct, some indirect). In addition, the causal and associative results (importance of factors) were different in magnitude and direction. This was the only study we found on surgical casual ML that conducted explicit causal discovery, i.e. determining the causal structure. The results highlight the disagreement between causal and associative models and higher ‘correctness’ of the causal models, hence the importance of incorporating causal inference.

Marafino et al. 2020 [8] (Kaiser Permanente) developed a causal machine learning framework (using causal forests) for predicting the most preventable hospital readmissions, as opposed to simply the highest risk of readmission as is done with predictive models. They looked at one treatment, a readmission programme. They predicted the impact of the treatment on specific patients, even if the treatment had not been applied yet. The model estimated how the treatment would change the outcome for that individual compared to if they had not received it. The patient cohort consisted of 1,584,902 hospitalisations at Kaiser Permanente’s Northern California hospitals from June 2010 to December 2018.

Marafino et al. showed that targeting interventions based on causal effect estimates could prevent more readmissions than using predicted risk alone. This can be viewed as a patient simulator to explore different scenarios, treatment, or no treatment. However, there is only one treatment, the readmission protocol, and this is not necessarily available in other centres.

Babayoff et al. 2022 [24] (Bar Ilan University, Sourasky Hospital) estimated ‘Duration of Surgery’ (DOS) using predictive models (DOSM), estimated influential factors using causal analysis and then built a predictor using variables with high causal effect (DOSM-F). The patient cohort consisted of 23,293 retrospective surgical records from the Tel Aviv Sourasky Medical Center, covering the eight most common surgeries between 2010 and 2020.

The authors found that most of the influential features of the predictive models do not have a causal relationship with DOS, corroborating the findings of Lee et al. [22]. The predictive model with causal features had slightly lower mean accuracy. They propose using the causal effect for clinical decisions. Including DOSM-F to estimate change in DOS as a result of changing causal feature values, similar to the scenario planning implemented by Marafino et al. [8]; however unlike Marafino et al. [8], they did not calculate individual treatment effects or have a single major modifiable treatment such as the readmission prevention program. They treated all variables as ‘treatments’, so the work is applicable to any centre, even if they do not have something like this readmission prevention program. But it’s on the population level, not the individual patient, so it cannot be used for decision making at the patient level.

These studies demonstrate the importance of causal modelling in surgical contexts. Lee et al. [22] showed it helps to have causal structure and Babayoff et al. [24] proposed a method to select features based on causal effect. Marafino et al. [8] identified preventable risks and allowed for forecasting different treatments, but it was designed around a readmission prevention program, which may not be applicable in other hospitals.

Nastl and Hardt [25] tested whether selecting causal features could improve generalisation of predictive models across ‘domains’, defined as subsets of samples containing feature and target variables. Domains were created by dividing datasets into subsets using selected variables. The researchers found that using all available variables provided a better trade-off between accuracy in-domain and accuracy out-of-domain. Their research overlaps ours in some ways, but there are some differences:

- The dataset we are using is specific to hospital surgeries, which is a domain not covered in their paper, and contains a broader set of variables, which may improve causal analysis.
- They identified causal and ‘plausibly causal’ features based on the literature and expert input, augmented by causal discovery. These feature groups are used as feature sets for training of predictive models, without adjustment for confounding or selection bias. We will adjust for bias, with the aim of training
- models on invariant relationships.
- We will first identify causal features using causal discovery and then verify them with domain experts. Identifying causal relationships supported by the data is required for the assumption that data adjustments could modify predictive performance.
- The hypertension dataset that they used was split into subsets using BMI, by separating patients who are overweight or obese from other patients. We split using weighted sampling to achieve overlapping populations that resemble a distribution shift.

Nastl and Hardt [25] suggest that further evidence is required to confirm whether causal techniques can in fact improve predictive performance; this study will test the techniques on a new dataset and different methodology.

## 4 Limitations

This study has several limitations due to the nature of the data and the methodology.

While simulating populations provides valuable insights, it may not fully capture the complexities of realworld differences. Future research should include two distinct environments to more accurately reflect diverse conditions and validate findings.

The causal discovery algorithms are limited in their ability to identify causal relationships, especially in cases of hidden confounders or colliders. To strengthen our approach, we will collaborate with clinicians from the hospital providing the data to validate our assumptions. However, the resulting causal graph is expected to provide only an approximate representation of the true data generation process. Consequently, controls limited only to variables identified in the causal graph may not address all causes of bias in the data, potentially leading to discrepancies in treatment effect estimates across different populations.

In addition, inaccuracies in the causal graph could flow on to CATE estimates, and in turn to the calculation of preventable risk. Nevertheless, we believe that this is a valuable exploration of the concept. Future work should include a prospective study where clinical procedures that can reduce risk are identified in advance and their impact in practice is compared with model estimates.

## 5 Data confidentiality

The INSPIRE dataset is a publicly available resource that has been de-identified by the data provider in accordance with relevant data protection standards. Investigators will access and use the dataset strictly under the terms and conditions set by the data custodian. No attempts will be made to re-identify any individual records or link them to other sources that could compromise confidentiality. All dataset files will be stored on a secure computer, accessible only to authorised study personnel. The research team will not share or distribute the raw data outside the parameters agreed upon with the data provider. Any results or publications derived from this research will present only aggregate findings and will not include any potentially identifiable information [26].

## 6 Waiver of consent

Because the INSPIRE dataset is a publicly available, de-identified dataset, no additional consent from individuals included in the dataset is necessary. The INSPIRE project on PhysioNet states: “This study was approved by the Institutional Review Board (IRB) of Seoul National University Hospital (SNUH, IRB No. H-2210-078-1368). The IRB also waived informed consent due to the retrospective nature of the study design. Additionally, the Institutional Data Review Board (DRB) of SNUH approved the release of the dataset to the public after a review of the dataset with the decision of adequate de-identification (BRB No. BD-R-2022-11-02).”

## Data Availability

The dataset is available online at https://physionet.org/content/inspire/1.3/

https://physionet.org/content/inspire/1.3/

